# The quorum sensing *com* system regulates pneumococcal colonisation and invasive disease in a pseudo-stratified airway tissue model

**DOI:** 10.1101/2021.12.16.21267943

**Authors:** Christian R. Kahlert, Susanne Nigg, Lucas Onder, Ronald Dijkman, Liliane Diener, Regulo Rodriguez, Pietro Vernazza, Volker Thiel, Jorge E. Vidal, Werner C. Albrich

## Abstract

*Streptococcus pneumoniae* (Spn) colonises respiratory epithelia but can also invade lung cells causing pneumonia. We developed an *ex vivo* model with human airway epithelial (HAE) cells harvested from lung biopsies to study Spn colonisation and translocation. Flow-cytometry, confocal imaging and electron microscopy studies identified the epithelial lineage with signs of differentiation (beating cilia, mucus, and tight junctions). HAE cells were challenged with Spn wild-type TIGR4 (*wtSpn*) or its isogenic Δ*comC* quorum sensing-deficient mutant. Δ*comC* mutant colonised significantly less than *wtSpn* at 6 h post-inoculation but at significantly higher levels at 19 h and 30 h. Translocation correlated inversely with colonisation density. Transepithelial electric resistance (TEER) decreased after pneumococcal infection and correlated with increased translocation for both strains.

Confocal imaging illustrated colocalisation of intracellular Spn with both cilia and zonulin-1 and prominent microcolony formation with wtSpn but disintegration of microcolony structures over time with Δ*comC* mutant. Δ*comC* caused a more pronounced release of both zonulin-1 and lactate dehydrogenase into the supernatant at later time points than wtSpn, suggesting that cytotoxicity is likely not the mechanism leading to translocation. There was a density- and time-dependent increase of inflammatory cytokines from human HAE cells infected with Δ*comC* compared with wtSpn, including increased levels of the NLRP3 inflammasome-related IL-18.

In conclusion, our experiments indicate that ComC system allows a higher organisational level of population structure resulting in microcolony formation, increased early colonisation and subsequent translocation. We propose that ComC inactivation unleashes a very different and possibly more virulent phenotype that merits further investigation.

## Introduction

Community-acquired pneumonia due to *Streptococcus pneumoniae* (Spn) is associated with high morbidity and mortality worldwide [1]. Currently there are at least 100 different pneumococcal serotypes known with different ability to cause mucosal and invasive pneumococcal disease (IPD) [2]. Colonisation with subsequent carriage in the upper airways is considered a “conditio sine qua non” for the development of pneumococcal respiratory tract infections and IPD [3, 4]. A critical colonisation density in the nasopharynx above which there is a high risk for pneumococcal pneumonia versus asymptomatic colonisation has been demonstrated in adults and children [5–7].

Spn encodes and produces different quorum sensing (QS) systems including the *com* QS system. Recent studies showed that Com regulates colonisation and biofilms production on human lung and pharyngeal cells [8]. Com, encoded by the locus *comABCD*, regulate expression of genes which are involved in competence-induced bacterial killing and fratricide, production of lantibiotics (Phr peptide QS), and increased virulence due to increased production of capsular polysaccharides [9].

To study pneumococcal-human interaction, animal models and immortalised cell culture models are generally utilised. The latter frequently consists of cancer cell lines, which represent an “artificial” environment lacking natural tissue diversity, differentiation and functionality (e.g. mucus production) of involved specialised airway cells and the 3D structure of the human lung epithelium, including the physiological air-liquid interface (ALI) and formation of tight-junctions (TJs)[10, 11]. Recently, an experimental human Spn carriage model has been developed [12][13, 14]. 24 h after individuals were infected with Spn serotype 6B strain, a different cytokine response was observed in those who were colonised by Spn compared to those infected but cleared the infection[15]. Whereas the human carriage model is helpful for carriage and vaccine studies, for ethical reasons pneumococcal virulence and lung pathogenesis can hardly be studied in human volunteers.

An *ex vivo* tissue model of primary human airway epithelial cells (HAE) with an air-liquid interface (ALI) was recently developed in our institution to study coronavirus pathogenesis[16]. This model includes a pseudo-stratified HAE cell layer containing basal, secretory, columnar, and ciliated cell populations. Thus, the *ex vivo* system recapitulates essential aspects of human airway epithelia, namely (i) presence of well-defined human airway epithelial cell types, which become (ii) functional and differentiated by producing mucus and showing ciliary activity, and (iii) maintains physico-biochemical barriers, such as the mucus layer, and a well-formed TJ belt with development of a trans-epithelial electrical resistance (TEER). These advantageous physiological characteristics and absence of an *ex vivo* model of human lung infection motivated us to adopt this *ex vivo* HAE-ALI model. We therefore infected differentiated human lung cells with a reference Spn strain TIGR4 (wildtype) or its isogenic Δ*comC* mutant and comprehensively compared colonisation, invasion, translocation, cytotoxicity, and human lung epithelial cell response.

## Material and methods

### Differentiated respiratory epithelium

Bronchial epithelial cells were isolated from bronchial biopsy samples of 5 different human donors and cryopreserved as previously described[16]. This was done in accordance with local regulations from Cantonal Hospital St. Gallen, Switzerland, St. Gallen Lung Biopsy Biobank (SGLBB) with approval by the ethics committee of the Canton St. Gallen (EKSG 11/044). Bronchial epithelial cells were cultured in flat bottom 12-well plates (Greiner bio-one, Switzerland) on an insert covered by a membrane with 1 µm pores until formation of a confluent differentiated epithelial monolayer with beating cilia and mucus production at the ALI apical side. Cells were counted to 500’000 per insert by flow cytometry (BD FACSCANTO II, Becton Dickinson, United States). This process was monitored by TEER measurement until a stable mean resistance was reached.

#### Pneumococcal infection

Spn strains were inoculated in a 200 µl suspension of balanced salt solution (BSS) on the apical side at different multiplicity of infection (MOI). ALI medium was added to the basolateral side. Pneumococci colonizing the apical compartment or invasive Spn in the basolateral compartment were counted by dilution and plating onto blood agar plates (measured as cfu/ml). Inactivated TIGR4 or latex particles served as negative controls.

***Further detailed methods on different bacterial strains, various outcome measures and statistical analysis are described in the supplement***.

## Results

### Primary HAE cells differentiate ex-vivo upon exposure to air

To model pneumococcal colonisation and invasive disease, cells from five different donors were cultured (Fig. 1). Integrity, and differentiation upon exposure to air [i.e., beating cilia (supplemental Movie 1-3), and mucus production] was observed by traditional *light microscopy* (Fig. 1). Confocal imaging showed ciliated structures and formation of a well-defined TJ complex, as evidenced by the presence of zonulin-1 (Fig. 2, ZO-1 in control cells). Ultrastructure of the TJ was revealed by *transmission electron microscopy* (supplemental Fig. 2A-C). We confirmed by *flow cytometry* that HAE cells were mostly (∼80%) epithelial cells [CD326+/CD31-, (supplemental Fig. 1B)]. Confocal microscopy confirmed the absence of fibroblasts (supplemental Fig. 1).**Thus, fully differentiated, pseudostratified, ciliated HAE cells are predominantly bronchial epithelial cells**.

**Figure 1.**
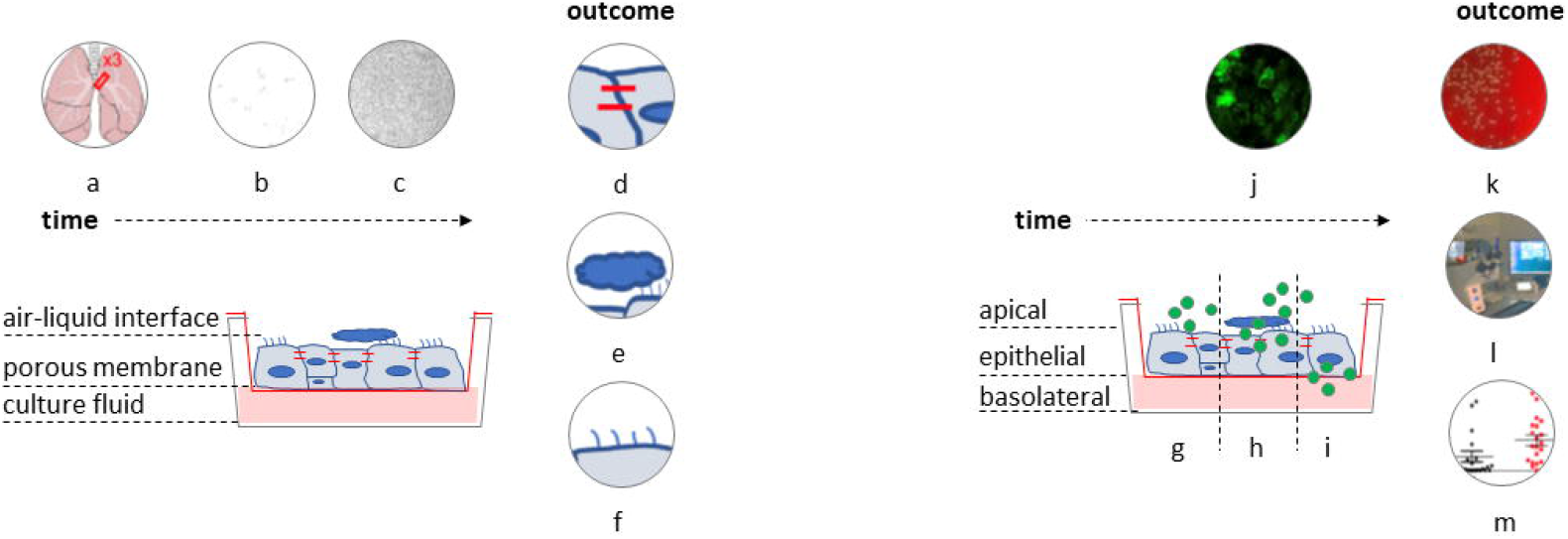
Modelling pneumococcal invasive disease: from colonisation (apical) to transmigration and translocation (basolateral) **Left, *non-infected model***. Primary human bronchial epithelial cells were isolated from bronchial biopsy samples from 5 donors (a). Cells were transferred as a single cell suspension (b) to a transwell insert that contains a membrane with pore size of 1 µm and incubated with culture medium in both the apical and basolateral side. Cells were incubated until the formation of a confluent monolayer when the cell culture medium was removed from the apical side to expose cells to air (air-liquid interface). Incubation was continued until the establishment of a differentiated pseudostratified bronchial epithelium (c) with tight junctions (d) mucus production (e), ciliary activity (f). **Right, *infected model***. To simulate the in-vivo pathogenesis and assess colonisation (g) and epithelial adhesion, transmigration (h) through the epithelium and finally translocation to the basolateral side (“invasive pneumococcal disease”, i), the established epithelium was inoculated on the apical side with different strains of *Streptococcus pneumoniae* (j) at 15 or 30 multiplicity of infection (MOI) and infected cells were incubated for 6h, 19h or 30h. Finally, 3 different spaces (apical, epithelial, basolateral) were evaluated as follows: Pneumococci in the apical and basolateral side were cultured (k), cytotoxicity of the epithelium was assessed by obtaining the transepithelial electrical resistance (TEER), the release of lactate dehydrogenase (LDH) as well as by confocal microscopy (l). In addition, the epithelial response to pneumococcus was evaluated for the release of cytokines and zonulin-1 (m) including the corresponding mRNA expression.

**Figure 2.**
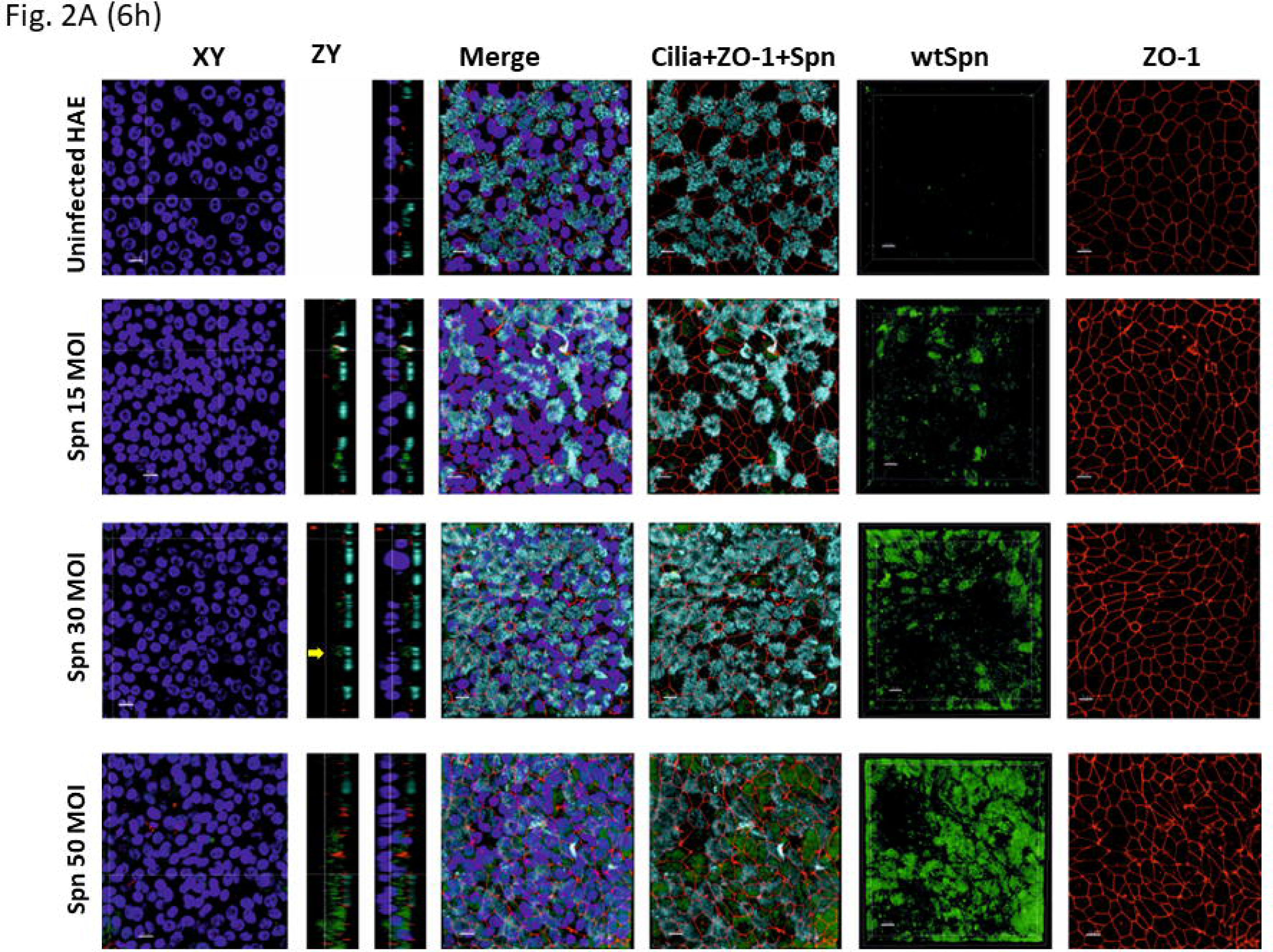
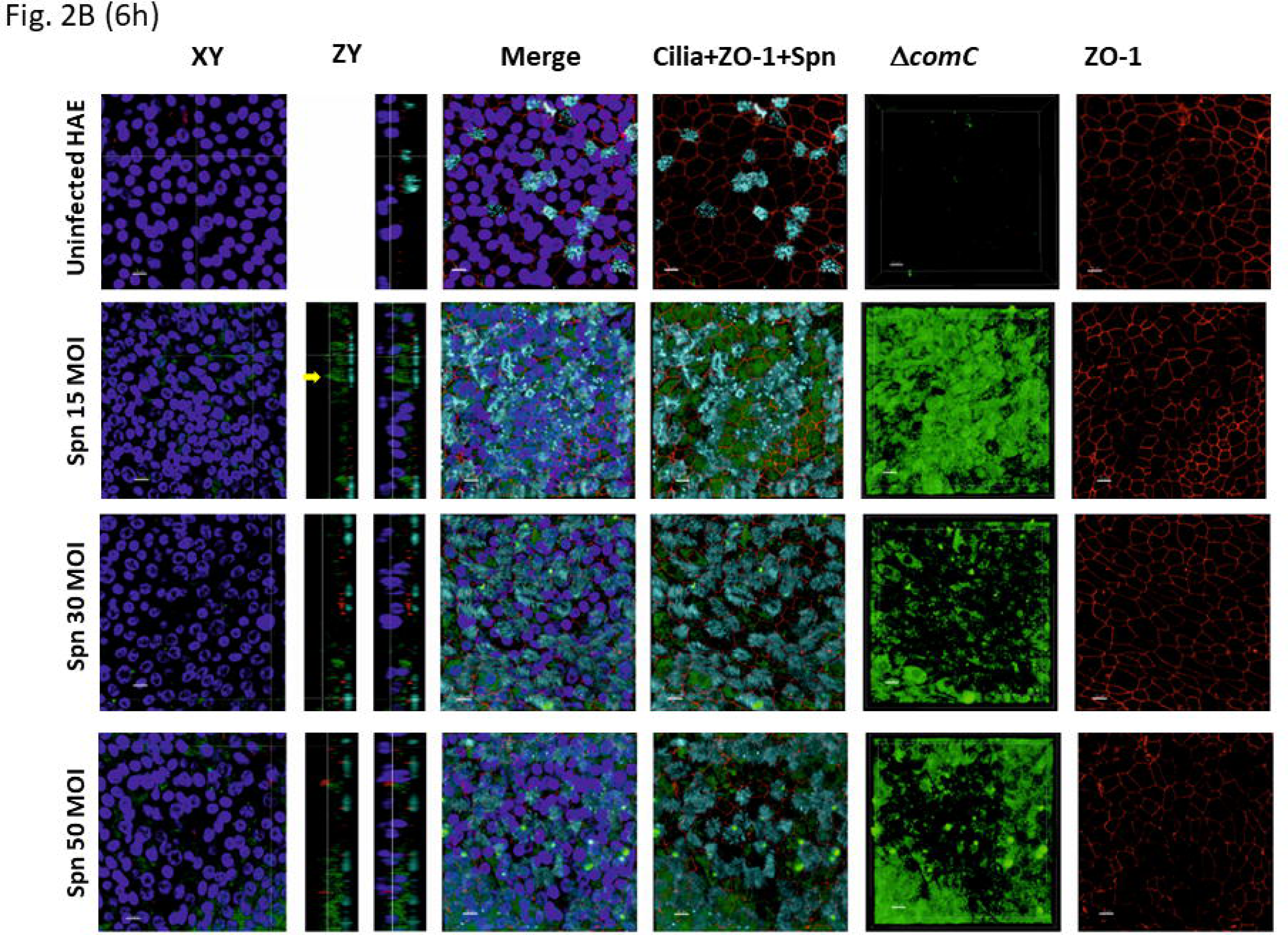
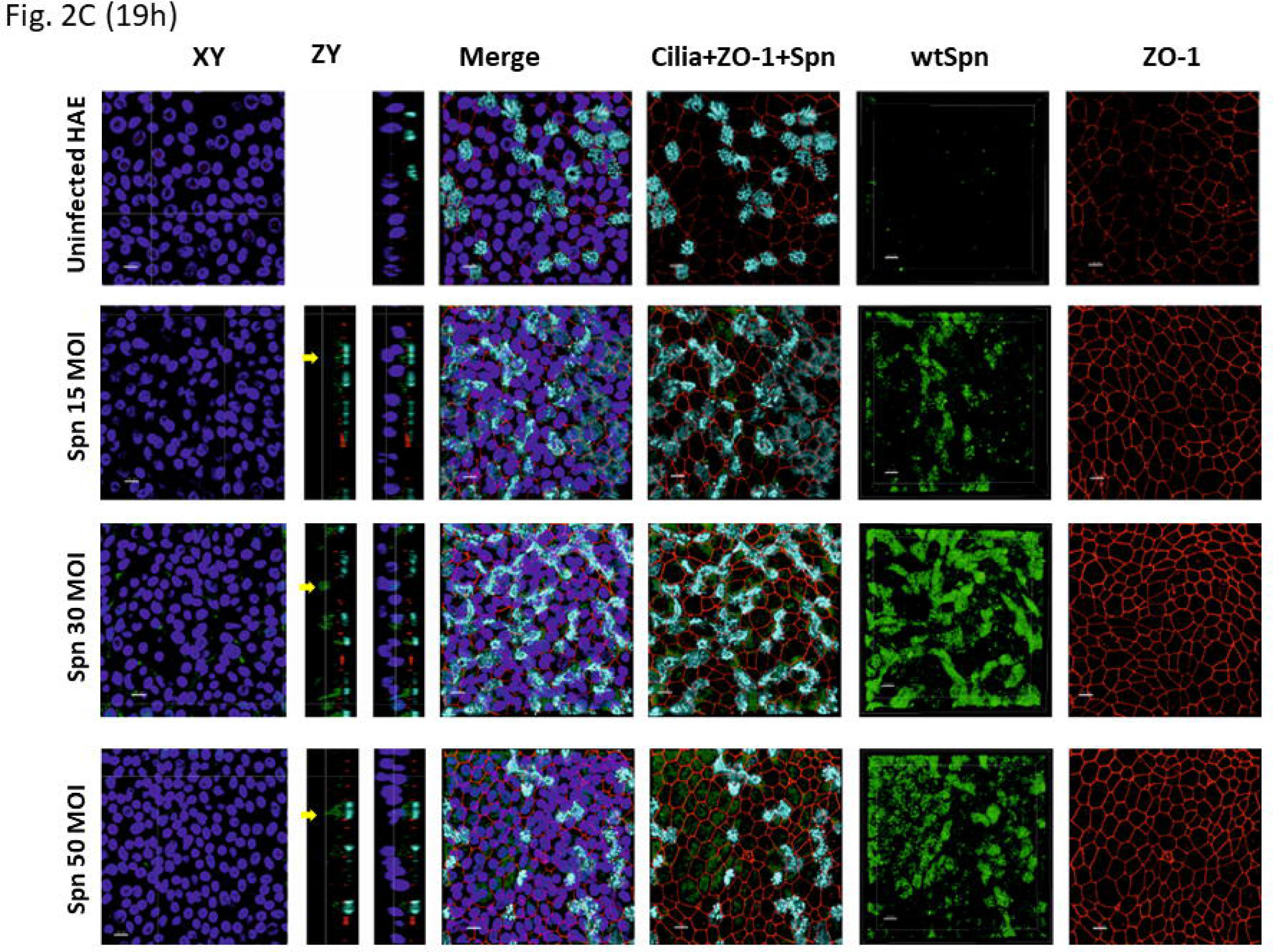
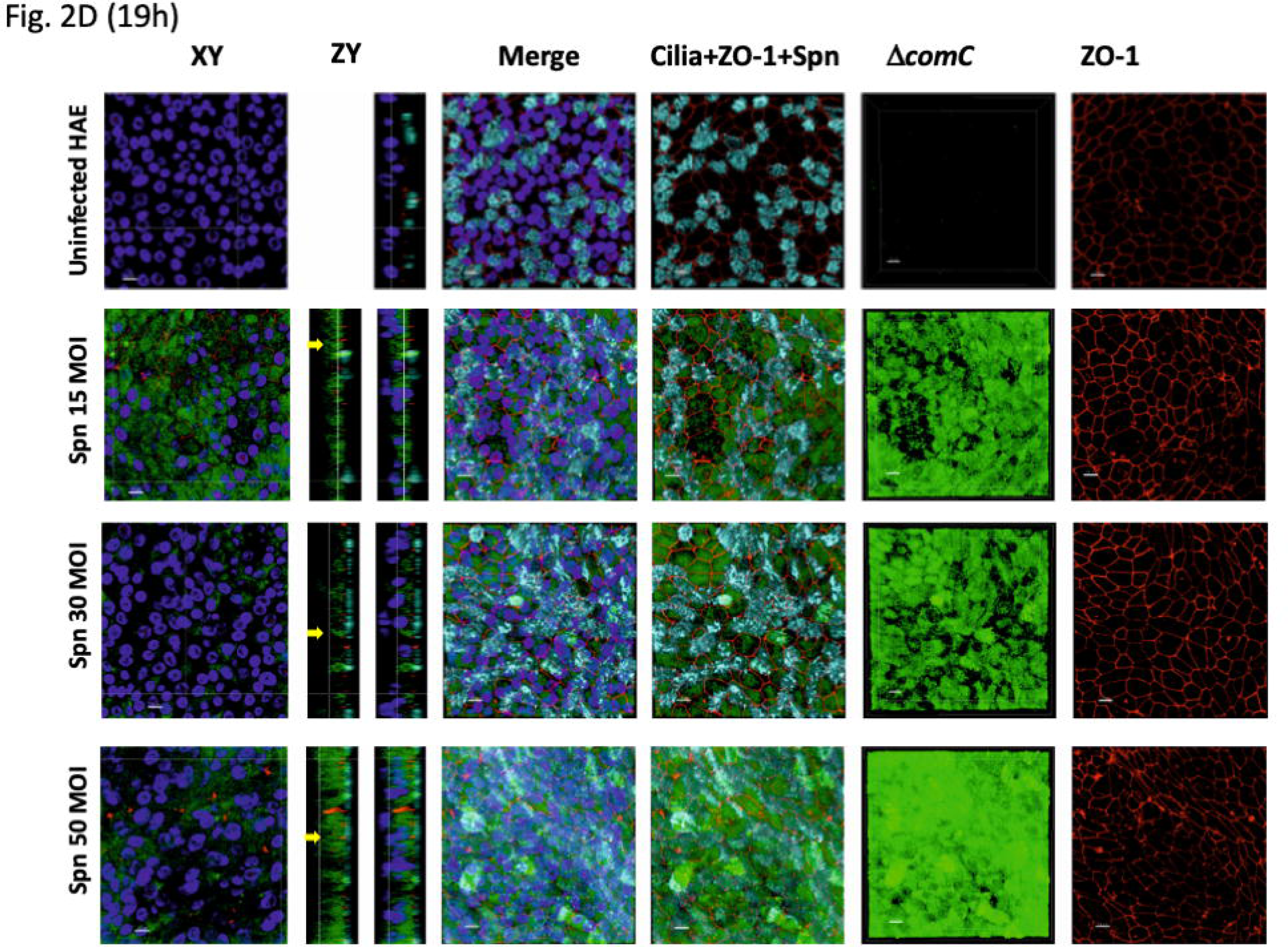
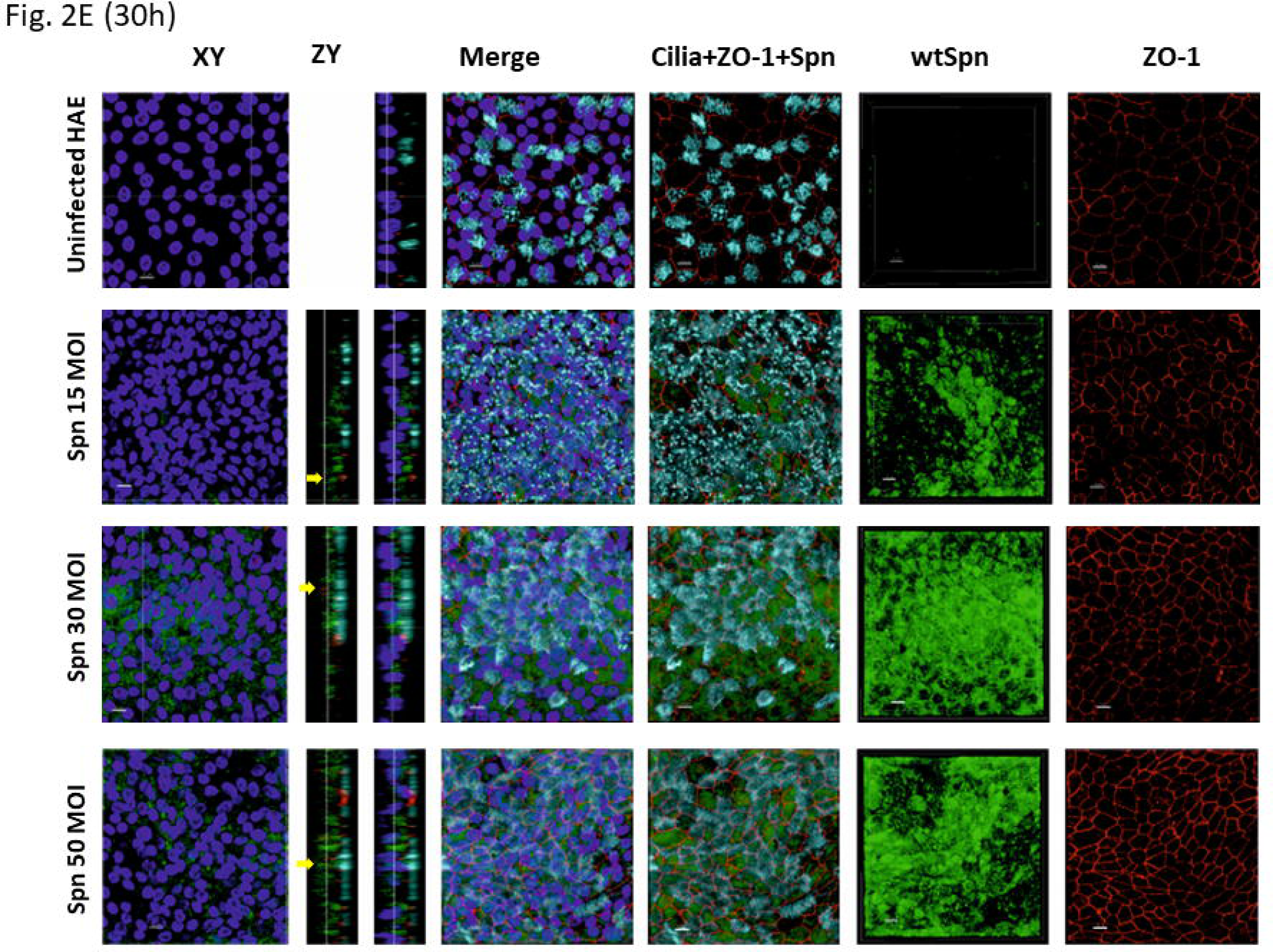
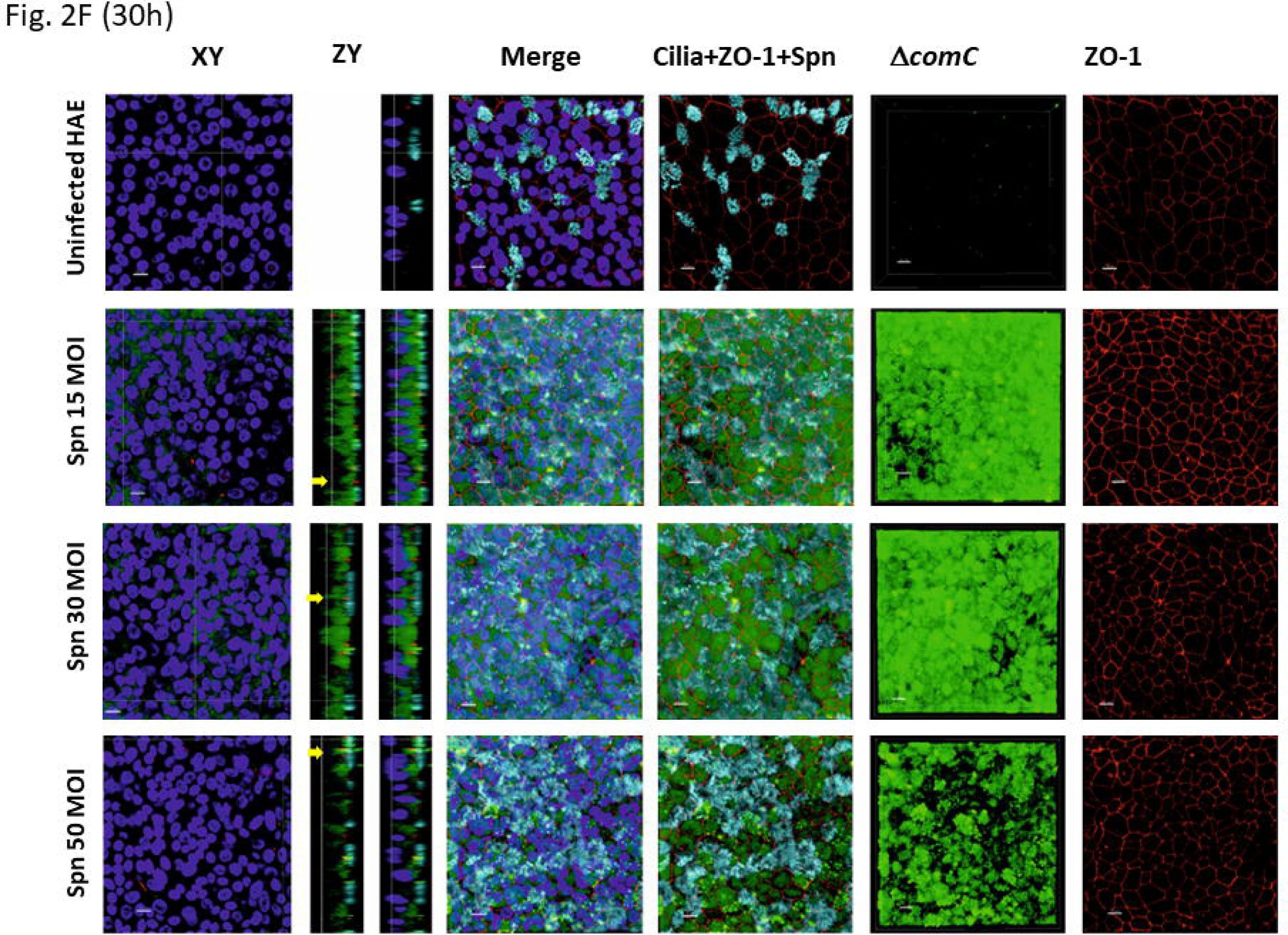
Confocal microscopy of human airway epithelial (HAE) cell cultures infected with wtSpn and ΔcomC mutant. HAE cells infected with wtSpn or Δ*comC* mutant are shown after 6 h (A, B), 19 h (C, D) and 30 h (E, F) post-inoculation. Within each panel (A-F), the top row shows uninfected HAE cells, the second row HAE cells infected with 15 MOI, the third row HAE cells infected with 30 MOI and the fourth row shows HAE cells infected with 50 MOI of pneumococci. Immunostaining, indicated on top of each panel, was performed with DAPI (4′,6-diamidino-2-phenylindole) for nuclei, monoclonal antibodies against anti-β-tubulin IV to detect ciliated cells (light blue), against ZO-1 for tight junctions (red) and fluorescein conjugated antibody against *S. pneumoniae* (green, indicated at TIGR4 or Δ*comC*). Columns depict stacks in XY plane or where indicated, orthogonal zy plane. Yellow arrow shows pneumococci underneath both cilia staining and ZO-1 signal. The preparations were examined with a confocal microscopy. Bars in all panels=10 μm.

### Inoculation of pneumococci results in stable apical colonisation at 6 hours

To resemble natural lung infection where nutrients are acquired by Spn from human cells, i.e., instead from the cell culture medium, BSS was added to the apical side of differentiated HAE cells before infecting the cells with wtSpn or the Δ*comC* mutant. Colonising Spn at the apical side reached the highest density (∼10^10^ cfu/ml) within 6 h of incubation after which there was no further increase (Fig. 3A, B, C). In contrast, low Spn frequency was cultured from the basolateral side (translocation, “invasive disease”) at this time point at any of the utilised inocula (Fig. 3D).

**Figure 3.**
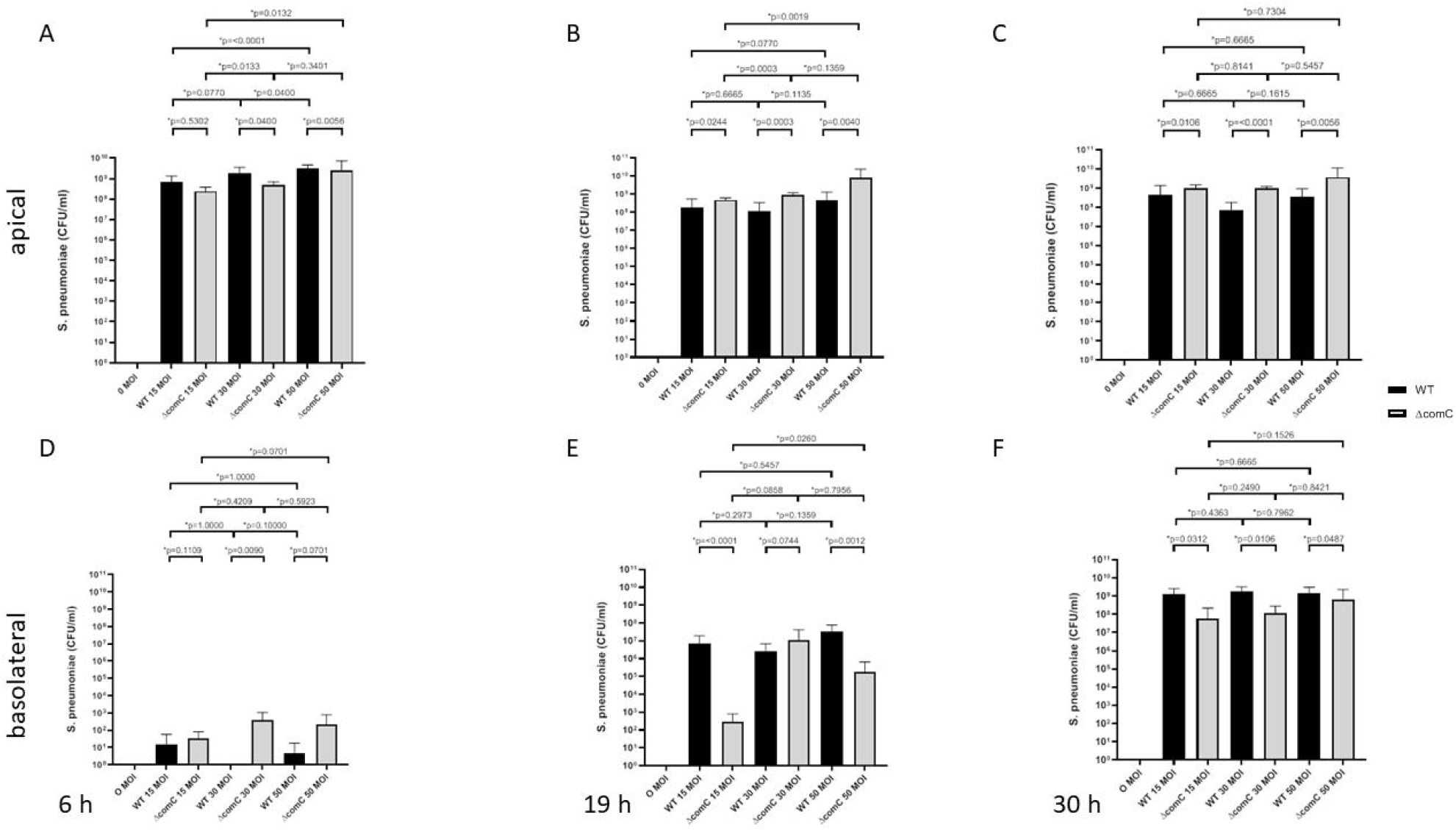
Quantification of Streptococcus pneumoniae colonisation of and translocation. Pneumococci were infected at the apical side of differentiated HAE cells at a multiplicity of infection (MOI) of 15, 30 and 50. The density of bacteria in the apical and basolateral space was obtained by dilution and plating (CFU) at 6 h, 19 h and 30 h post-inoculation.

Three-dimensional (3D) reconstruction of confocal z-stacks and analysis of orthogonal zy planes demonstrated Spn on the apical side of differentiated HAE cells (i.e., above the TJ staining) (Fig. 2A, arrow). Spn formed microcolonies particularly colocalising with cilia (Fig. 2A, 2B, arrow). Dotted cilia were noticed in areas where Spn colonised HAE cells suggesting that bacteria had degraded cilia (Fig. 2 A-F, arrows). **Taken together, this suggests that at 6 h post-inoculation, pneumococci were mainly attached on top of HAE cells, representing colonisation**.

### Loss of *com* quorum sensing induces a disorganised adhesion phenotype with translocation to the basolateral side

An important role of the Com QS system is to provide extracellular DNA (eDNA) during colonisation, a response mediated by a density threshold. We therefore assessed the adhesion phenotype and translocation of the isogenic Δ*comC* mutant strain. At 19 h post-inoculation there was a trend for increasing apical Spn densities with increasing doses whereas this trend was not seen after 30 h suggesting steady state of colonisation after 6 h (Fig 3 A-C). Compared to wtSpn, the Δ*comC* colonised significantly less at 6 h post-inoculation but at higher levels at 19 h and 30 h (Fig. 3 A-C). In contrast, more Δ*comC* than wtSpn bacteria translocated at 6 h. Compared to 6 h, there was an increased number of Spn identified at the basolateral side at 19 h with higher counts for wtSpn than Δ*comC* mutant (Fig. 3 D-F). There was a further increase in translocation for both strains at 30 h with higher counts for wtSpn (Fig. 3 E, F).

In confocal imaging, there was prominent microcolony formation with wtSpn on top of HAE cells at 19 h and 30 h compared to increasing disintegration of microcolony structures over time with Δ*comC* (Fig. 2 C-F). zy confocal sections revealed the colocalisation of intracellular Spn with both cilia and ZO-1 at all time points (transmigration, Fig 2 C-F, yellow arrows). **These experiments suggest that pneumococcus primarily attaches to ciliated structures to gain access to human lung cells (Fig. 2A, 2B, yellow arrows). Translocation to the basolateral side increased from 6 h to 19 h to 30 h post-inoculation correlating inversely with colonisation density**.

### Pneumococcal translocation correlates with a decreased transepithelial electrical resistance, that is specific and depends on time and dose of Spn

To begin studying the invasion mechanism, TEER was measured across HAE cells infected with TIGR4 as a correlate of the integrity of the TJ complex (Fig. 4A). Overall, a similar trend of decline was present over time, but with varying magnitude and timing. Starting TEERs ranged from 600-800 Ω/cm^2^. There was a drop of median TEER at 6 h post-infection followed by a continued decrease. At 19 h post-infection, TEER had decreased by ∼60% (Fig. 4A) and by ∼85% at 28 h post-inoculation in comparison to the uninfected control (Fig. 4B). A similar time-dependent TEER decrease was observed when HAE cells were inoculated with a lower dose (Fig. 4B) or with another reference Spn strain (ATCC 49619, Fig. 4C). However, this decrease was specific for pneumococcal infection as evidenced by a similar TEER course after incubation with latex particles (Fig. 4C) or with heat-killed wtSpn (supplemental Fig 3) or BSS alone (0 MOI). Increasing inocula were associated with faster TEER decrease in wtSpn (r –0.81, p=0.002; Fig. 4C). A time-course study revealed that TEER decreased earlier and more rapidly with the Δ*comC* mutant compared to wtSpn while levelling off at 19 h and at 30 h with different slopes for different inocula. The effect of the strain on TEER showed a significant interaction with time (p<0.001) with faster decrease with Δ*comC* with a significant TEER differences at 6 h but not at 19 h and 30 h (p=0.01, p=0.18 and p=0.15, respectively; Fig. 4D). **Thus, periods of faster TEER decrease correlated with increased translocation**.

**Figure 4.**
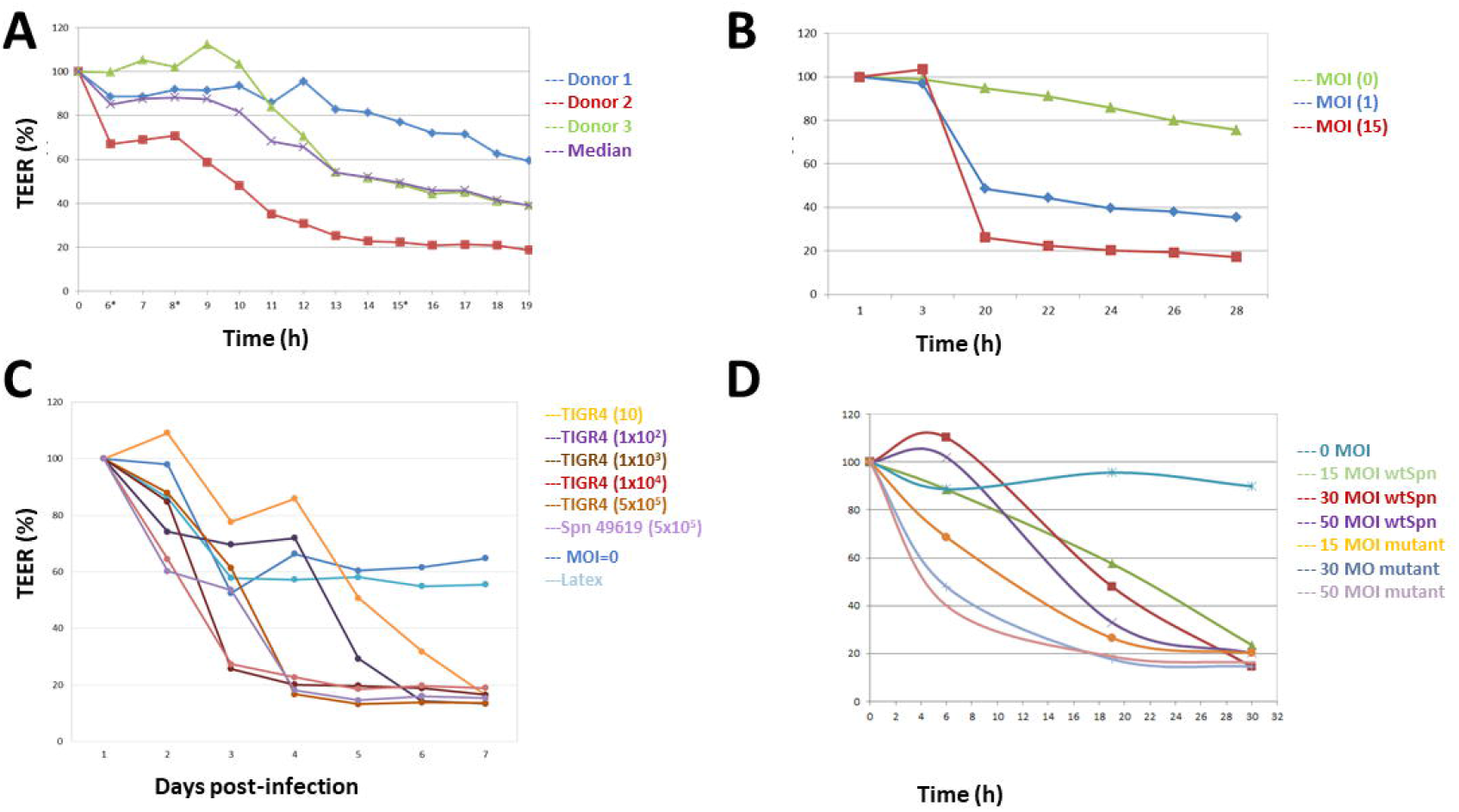
Infection with Streptococcus pneumoniae decreases the transepithelial electrical resistance of differentiated HAE cells. Differentiated HAE cells were infected with wtSpn at 15 MOI from 3 donors. The transepithelial electrical resistance (TEER) was obtained every hour between 6 h through 19 h post-infection (A). HAE cells were infected with wtSpn at 0, 1, or 15 MOI and TEER was evaluated at the indicated time points (B). HAE cells were infected with different pneumococal strains (wt-TIGR4, ATCC 49619) at different concentrations or mock infected with buffer or latex particles. TEER was evaluated every day (i.e., 24 h) post-infection for 7 days (C). TEER was measured at different timepoints post-infection with wtSpn or TIGR4 Δ*comC* mutant at different concentrations (D). Baseline TEER prior to infection was set to 100%. Changes of TEER are shown as percentage of decrease considering the baseline TEER maximum (100%) resistance (A-D).

### Pneumococcal translocation is not induced by cytotoxicity

To evaluate whether translocation of pneumococci was due to cytotoxicity lactate dehydrogenase (LDH) release in the supernatant was measured after inoculation of HAE cells with wtSpn or Δ*comC*. LDH levels were similar 6 h post-inoculation with both strains but significantly higher with Δ*comC* after 19 h and 30 h of incubation (Fig. 5A). For both wtSpn and Δ*comC*, LDH levels significantly increased with longer incubation (Fig. 5A). Higher LDH levels were observed with Δ*comC* mutant compared to wtSpn for all inoculation doses (Fig. 5B). When there was no translocation, LDH levels did not differ between both strains until 19 h but were higher with Δ*comC* after 30 h (Fig. 5C). In contrast, with translocation, higher LDH levels were seen with Δ*comC* compared to wtSpn at 19 h and 30 h (Fig. 5D). Surprisingly, among wtSpn, inserts with translocation showed significantly lower LDH levels at 19 h and 30 h than those without translocation (Fig. 5E). In contrast, among Δ*comC*, LDH levels were similar regardless of translocation (Fig. 5F).

**Figure 5.**
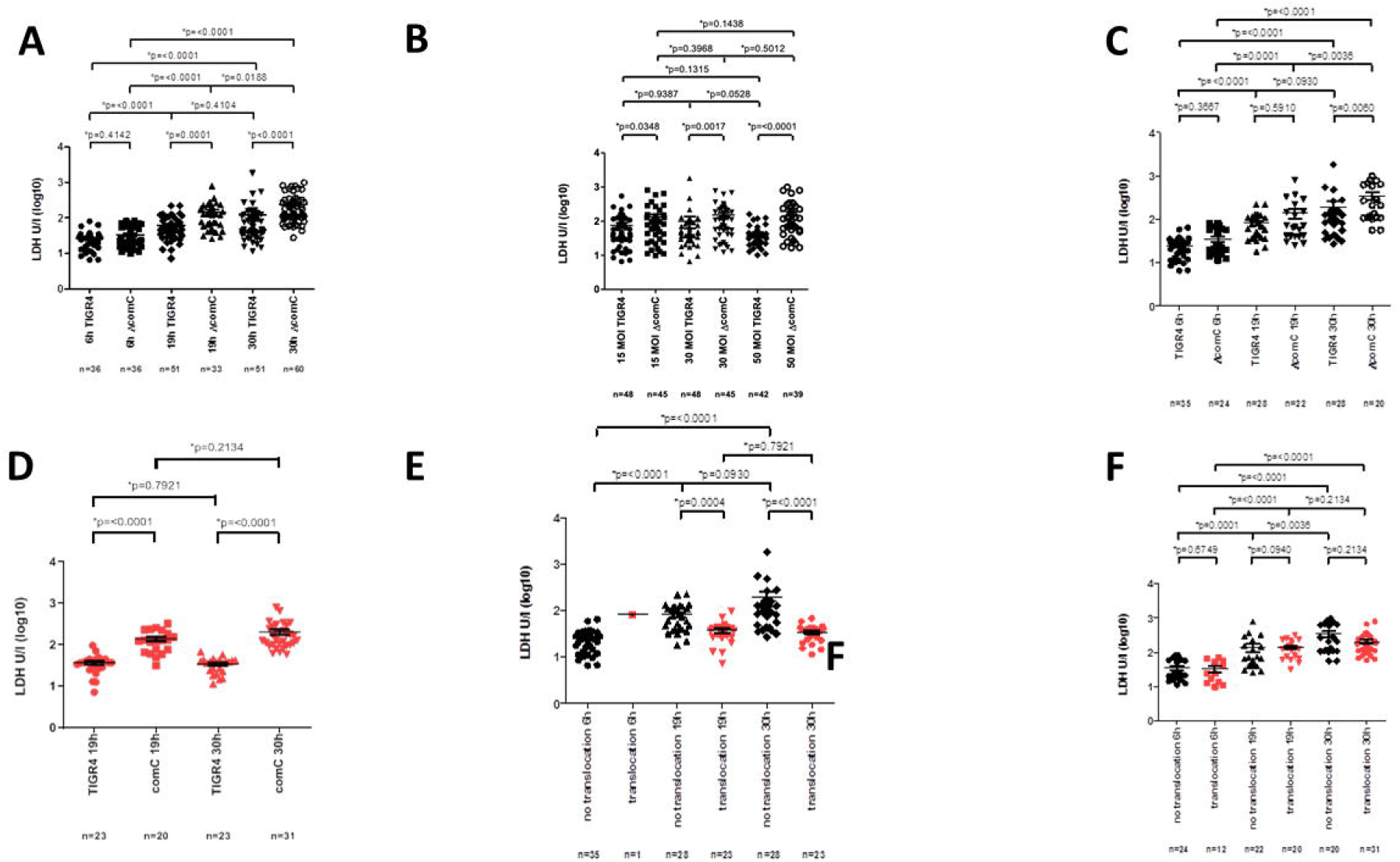
The absence of the comC system results in increased cytotoxicity. Lactate dehydrogenase (LDH) release was quantified (U/l) in supernatants of differentiated HAE cells, harvested from three different donors and inoculated with wtSpn or Δ*comC* mutant. LDH release after infection for different time points (A) and different concentrations (MOI) of wtSpn and Δ*comC* (B). LDH release is shown for infection with both strains in inserts without (C) and with (D) translocation at different time-points. LDH release from cells is shown for inserts without and with translocation from HAE cells infected with (E) wtSpn and Δ*comC* mutant (F). Error bars represent the standard errors of the means calculated using data from five independent experiments, that used HAE cells from three different donors and that included several transwell inserts in each biological replicate. Inserts with translocated bacteria are indicated in red, inserts without translocated bacteria in black (panel C-F). The total number (n) of transwell inserts with differentiated HAE cells tested is indicated under each strain/condition. *p*-values are shown on top of each comparison. *Indicates statistical significance *p*<0.05.

Spn also produces a homolog LDH enzyme during its metabolism [17]. We therefore quantified the amount of LDH in cultures of wtSpn grown for up to 30 h and LDH levels were below the limit of detection (15 U/L).

**Collectively, this indicates increased cytotoxicity by infection with** Δ***comC* mutant than wtSpn. However, cytotoxicity is likely not the mechanism leading to translocation**.

### Inoculation of HAE cells with pneumococci results in disintegration of the TJ

Since intracellular pneumococci colocalised with ZO-1 staining, we quantified the release of ZO-1 into the supernatants using ELISA. ZO-1 levels were significantly higher in HAE cells infected with Δ*comC* mutant than wtSpn (p<0.0001; Fig. 6A, 6B). ZO-1 levels increased over time, particularly in HAE cells infected with Δ*comC* (Fig. 6B) but were independent of inoculum density (Fig. 6C) and were not linked to translocation (Fig. 6D).

**Figure 6.**
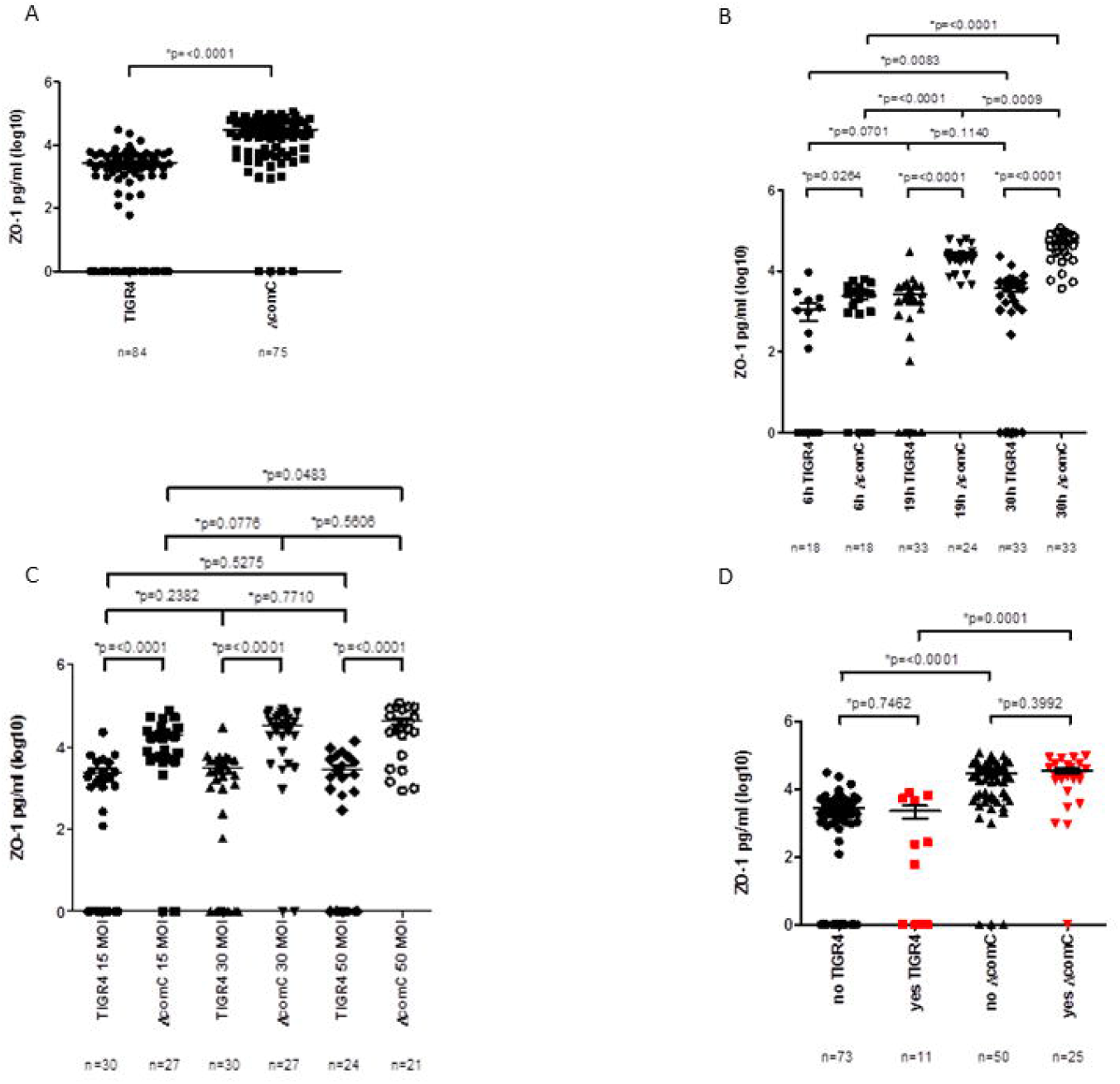
Detection of ZO-1 in the supernatant is higher in ΔcomC mutant, increases with time and is independent from translocation and dose. ZO-1 release was quantified in supernatants of differentiated HAE cells harvested from two donors with wtSpn and Δ*comC* mutant at different concentrations (15, 30, or 50 MOI) and different time points (6 h, 19 h and 30 h). ZO-1 levels are depicted in pg/ml after infection of HAE cells with wtSpn and Δ*comC* mutant overall (A), for different time points (B) and concentrations (C) and stratified for both strains each without translocation (“no”) and with translocation (“yes”) (D).

To investigate whether ZO-1 in the supernatant was released from damaged HAE cells or resulted from upregulated gene expression, ZO-1 mRNA was measured in infected HAE cells. Lower ZO-1 transcripts were detected in HAE cells infected with Δ*comC* mutant than with wtSpn (Fig. 7A). This became apparent at 19 h and was significant at 30 h post-infection (Fig. 7B), but without a dose-response effect (Fig. 7C). There was a non-significant trend for downregulation of ZO-1 expression in inserts with translocation compared to those without translocation, both for wtSpn and Δ*comC* (Fig. 7). **Overall, these results confirm that increased ZO-1 levels in supernatant cannot be attributed to increased ZO-1 expression but rather to increased release of ZO-1 into the supernatant**.

**Figure 7.**
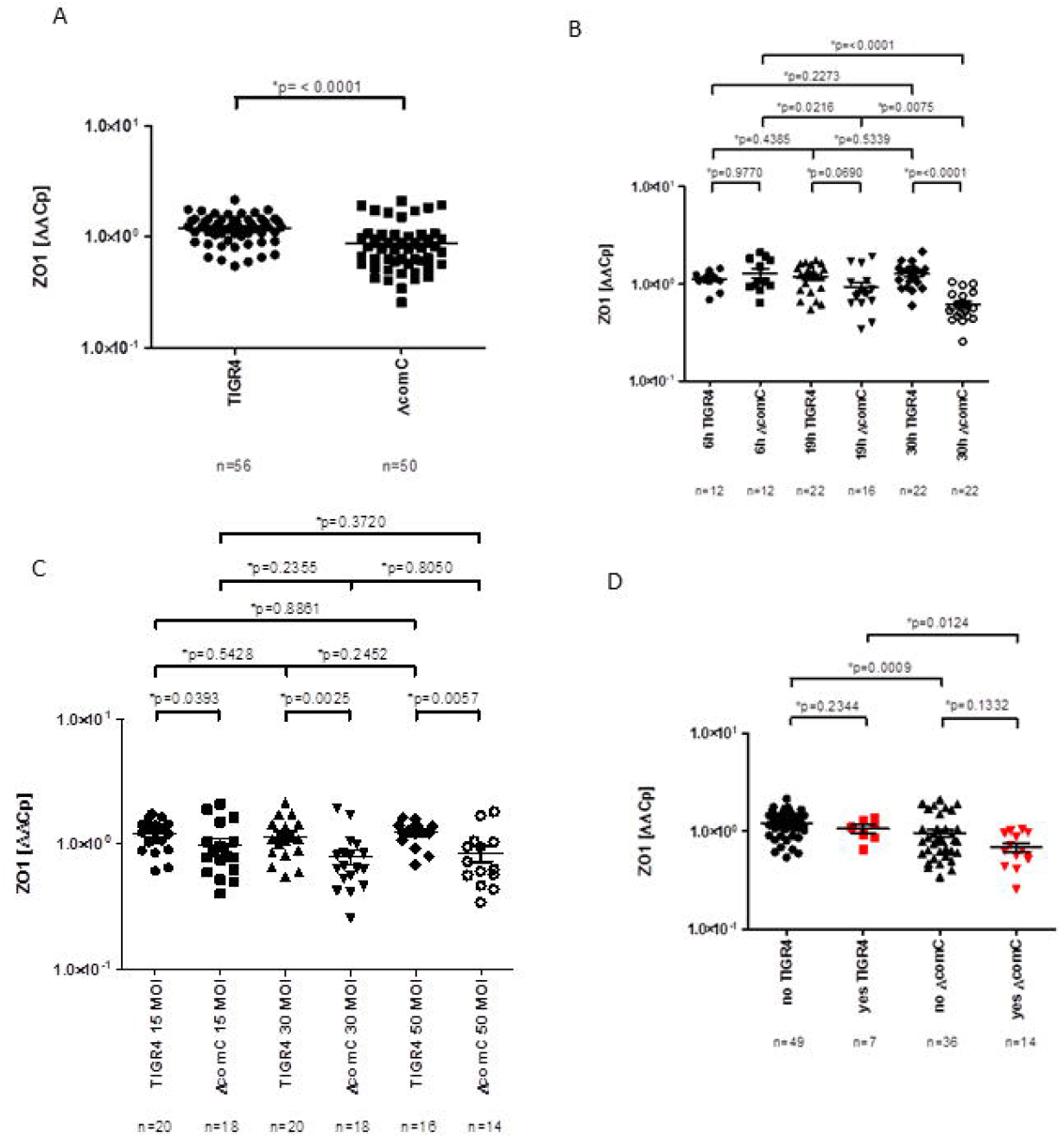
Detection of mRNA expression of the ZO-1 gene is lower in HAE ΔcomC mutant compared to wtSpn. mRNA expression of the ZO1-gene was measured in differentiated HAE cells infected with wtSpn or ΔcomC mutant at different concentrations (15, 30, or 50 MOI) and at different time points (6 h, 19 h and 30 h). ZO-1 mRNA expression is depicted as fold changes relative to uninfected control HAE cells and adjusted using mRNA expression of the gene encoding β-actin overall (A), for different time points (B) and concentrations (C) and stratified for both strains each without translocation (“no”) and with translocation (“yes”) (D).

### Temporal expression of specific host cytokines reflects colonisation and translocation

Secretion of 42 different human cytokines was evaluated during colonisation and translocation in the supernatants of HAE cells inoculated with pneumococci in comparison to uninfected differentiated HAE cells control. Remarkably, compared to non-infected cells, at the initial colonisation stage only IL-18 was significantly increased ∼60-fold in HAE cells infected with wtSpn and Δ*comC* mutant (Fig. 8A-8E). Furthermore, in HAE cells infected with the Δ*comC*, levels of IL-18 were significantly increased at 19 h and 30 h post-infection (Fig. 8B and 8C) compared to the 6 h colonisation stage, whereas there was no further increase for wtSpn beyond 6 h (Fig. 8B and 8C).

**Figure 8.**
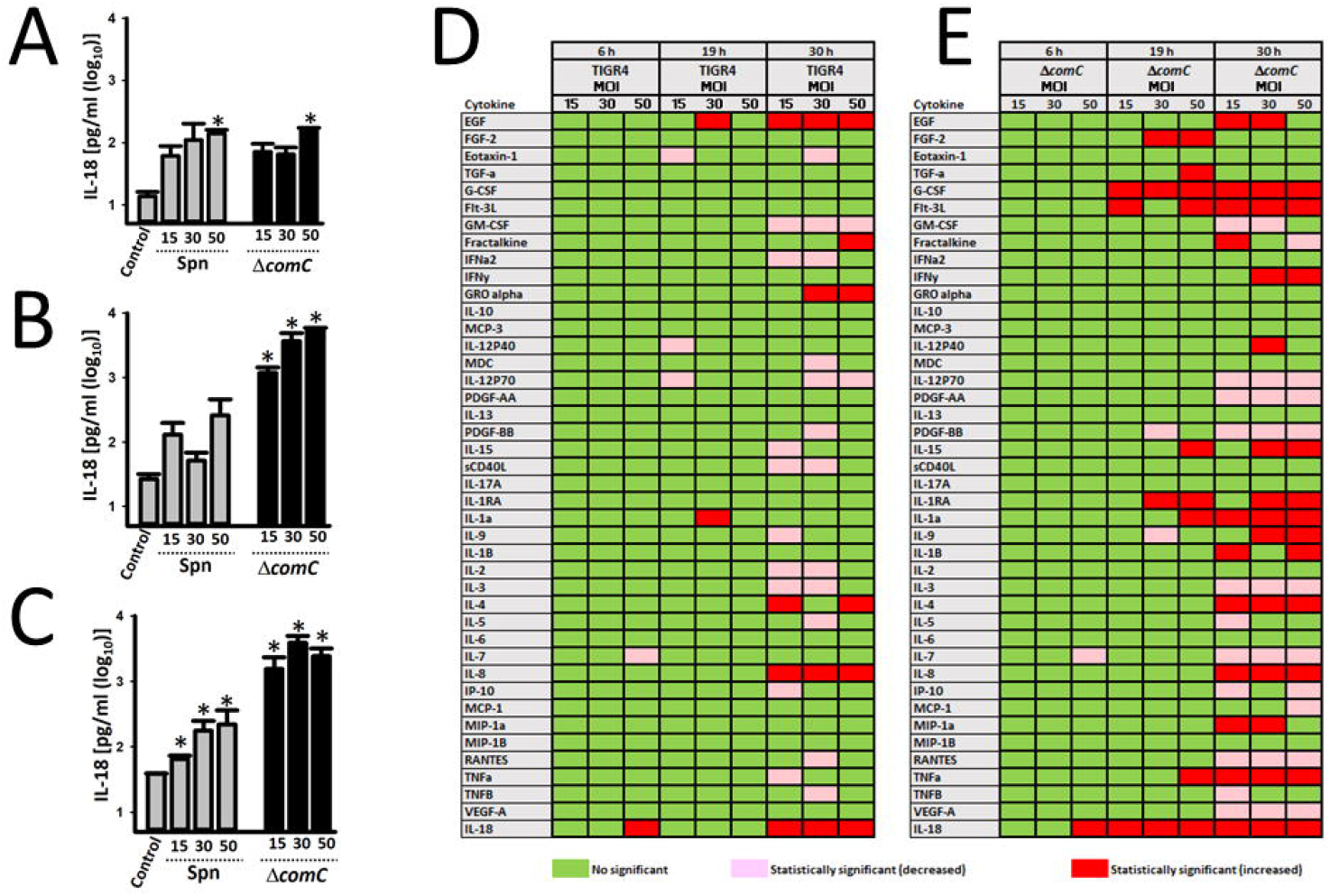
Cytokine release into the supernatant correlated with increased cytotoxicity post pneumococcal infection in ΔcomC mutant. Cytokines levels were measured in supernatant from HAE cells with different multiplicity of infection (MOI=15, 30, or 50) of wtSpn or ΔcomC mutant and incubated for 6 h, 19 h, or 30 h. IL-18 levels post-infection are shown at 6 h (A), 19 h (B) or 30 h (C). *p<0.05 refers to statistical significance versus IL-18 levels of supernatants of the uninfected control. Color coding of boxes indicates significantly (p<0.05) increased (red) or decreased (pink) of the cytokine levels in the supernatant with wtSpn (D) or ΔcomC mutant (E) compared to the uninfected control. Green boxes refer to similar cytokine level as compared to the uninfected control.

Infection with wtSpn did not consistently induce a significant change in the secretion of other cytokines at 6 h and 19 h, in comparison to the uninfected control. Only at 30 h post-infection, secretion of IL-8 and epidermal growth factor was significantly increased at all inoculated densities (Fig. 8D). In contrast, differentiated HAE cells infected with Δ*comC* mutant had significantly higher levels of G-CSF at 19 h post-infection and at 30 h increased levels of IL-1α, IL-1RA, IL-1β, IL-4, IL-8, IL-9, TNF-α, and significantly decreased detection of several cytokines including IL-3, IL-7 and RANTES (Fig. 8E). **Together, our data reveal an increased cytotoxic phenotype upon inactivation of *com* quorum sensing signalling**.

## Discussion

This differentiated HAE model simulates the entire pathway from pneumococcal colonisation of human lower airways to transepithelial migration and IPD. Cultures from the basolateral space revealed an infectious dose- and time-dependent penetration through the respiratory epithelium accompanied by an epithelial response. Confocal imaging demonstrated co-localisation of pneumococci with TJ and cilia and subsequent degradation of ciliary and TJ structures. Translocation into the basolateral space coincided with loss of epithelial integrity as shown by a TEER decrease. The host response to pneumococcal invasion was characterised by an increase of proinflammatory cytokines but it was altered when the *com* QS system was inactivated. Our model suggests that Com is associated with microcolony formation during colonisation and translocation, whereas inactivation of *comC*-mediated signalling correlates with epithelial disruption with increased cytotoxicity, and an increased inflammatory response.

The Com pathway is a well-studied QS system. It allows bacterial intercellular communication and enables fratricide, uptake of exogenous DNA and genetic recombination [8, 18]. *comC* encodes the competence stimulating peptide (CSP) that activates the QS system. Its inactivation results *in vitro* in a decreased production of early stages of biofilms[8, 19].

Likewise, in the current study, inactivation of *comC* led to early reduced and disorganised pneumococcal colonisation along with increased cytotoxicity and upregulated cytokine response but impaired transmigration. This is consistent with loss of a highly orchestrated host-pathogen interaction. Microinvasion was followed by an inflammatory response which was postulated as a mechanism for subsequent pneumococcal clearance[20]. Given the increased identification of hormones and molecules that block bacterial QS systems, it is likely that blocking *com*-QS may modulate severe cases of human pneumococcal disease. This potential mechanism is under investigation in our laboratories.

Upregulation of proinflammatory cytokines correlated with cytotoxicity. For example, levels of both IL-18 and LDH were increased in those HAE cells infected with Δ*comC* mutant compared to those infected with the wtSpn. The release of activated IL-18 and IL-1β is a hallmark of canonical - caspase-1-dependent-activation of the NLRP3 inflammasome[23]. Cell death can occur by uncontrolled activation of the NLRP3 inflammasome, known as pyroptosis[24, 25]. IL-18 has proinflammatory activity both IFNγ-dependent and -independent[26]. Therefore, evidence from this model indicates that pneumococci elicit a distinct IL-18 mediated inflammatory response that can induce differentiated human lung cells to undergo pyroptosis, particularly when *com*-QS is inactivated.

Except for immortalised cell cultures and animal models, we are aware of only two published lung tissue models [20, 27]. One model investigated infection on fresh lung tissue cylinders. In contrast, in our model bronchial epithelial cells are harvested to a single cell suspension that allows cryopreservation and stocking cells for repeated experiments. The other model describes an experimental human pneumococcal carriage model and in addition a HAE model that recapitulated in vivo findings [20]. This epithelial model is very similar to ours. Importantly and confirming the applicability of our HAE model to *in vivo* situations, Weight et al. also reported adhesion, microcolony formation and epithelial invasion of a serotype 6B pneumococcus. This model focussed on the first 3 hours of pneumococcal infection whereas we studied infection for up to 30 hours.

There are several major strengths of our experimental setup which make it more physiologic compared to traditional cell cultures. First, we used a more realistic *ex vivo* model with differentiated cells. Secondly, pneumococci were inoculated on the apical side in the absence of cell culture medium. Consequently, bacteria acquired nutrients directly from their interaction with differentiated HAE cells. Unlike homogenous colonisation on the cell substrate seen in models where bacteria are inoculated in cell culture medium, in our model, colonising pneumococci formed microcolonies. Similarly, Trappetti et al. also reported that a wtSpn strain formed microcolonies whereas the isogenic Δ*comC* mutant was unable to attach[19]. Our *ex vivo* model indicated a loss of microcolony formation over time with a disorganised adhesion pattern in those infected with an inactivated *comC* system. Third, epithelium and response to infection is donor specific. Results that are generally reproducible with different human donors can thus be generalised. Donor-specificity is a great asset as this model reflects the individual physiological situation and susceptibility to colonisation and infection more closely than artificial systems or immortalised cell lines. Limitations of our HEA model include the lack of an adaptive immune response and the time required for epithelial differentiation.

In conclusion, our experiments indicate that the *com*C system allows a higher organisational level of population structure resulting in microcolony formation, increased early colonisation and subsequent translocation. We propose that inactivation of QS unleashes a very different and possibly more virulent phenotype of pneumococci that merits further investigation. This finding was observed using an *ex vivo* primary HAE model that recapitulates pathophysiological events of pneumococcal colonisation and invasion. It may thus allow identification of novel mechanisms to improve diagnostics and determine new preventive and therapeutic targets to combat pneumococcal diseases.

## Data Availability

All data produced in the present study are available upon reasonable request to the authors

## Acknowledgement

We thank Sabine Güsewell, PhD sincerely for the great expert statistical advice.

## Contribution

CRK and WCA initiated the study. CRK, WCA and JEV developed the design and methods. SN performed all experiments with exception of transmission electron microscopy, which was performed by LD. LO supervised confocal microscopy. CRK, SN, JEV and WCA performed statistical analyses. CRK, JEV and WCA wrote the manuscript. CRK, WCA, JEV, SN and LO contributed to data acquisition and interpretation. All authors revised and approved the manuscript for intellectual content. CRK, JEV and WCA had full access to the data in the study, all authors agreed to the final version including submission for publication and accept responsibility for this work.

## Funding

Work in the Vidal laboratory was in part supported by grants from the National Institutes of Health (NIH; 1R21AI144571-01, and 1R21AI151571-01A1).

Work at the experimental infectiology laboratory at the Cantonal Hospital of St. Gallen was supported by the Lungenliga St. Gallen, Medical Research Centre, Cantonal Hospital St.Gallen, funds of the Division of Infectious Diseases & Hospital Epidemiology, Cantonal Hospital St.Gallen.

## Notes

### Competing Interest Statement

WA: attendance at advisory boards for Pfizer, MSD, Sanofi, all paid to his institution

### Author Declarations

EKOS (Ethikkommission Ostschweiz), EKSG 11/044

