## Supplementary figures and images for "The quorum sensing *com* system regulates pneumococcal colonisation and invasive disease in a pseudo-stratified airway tissue model"

### Fig. S1A

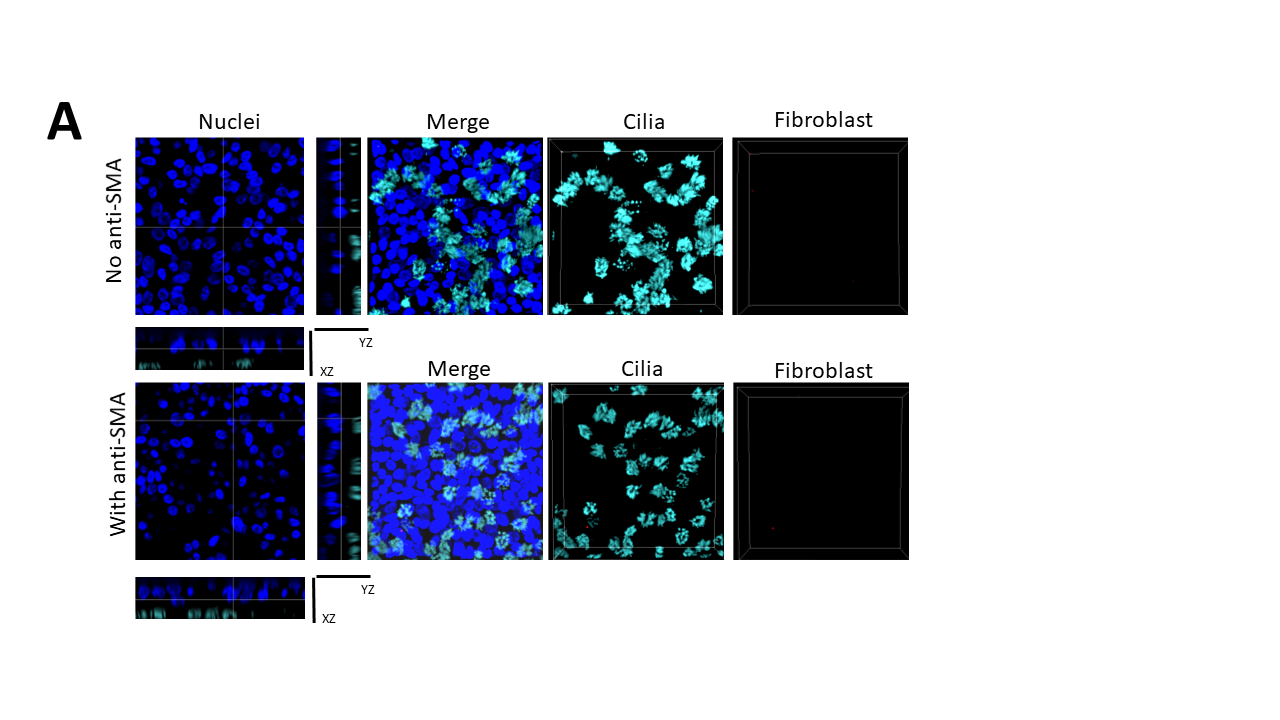

### Fig. S1B

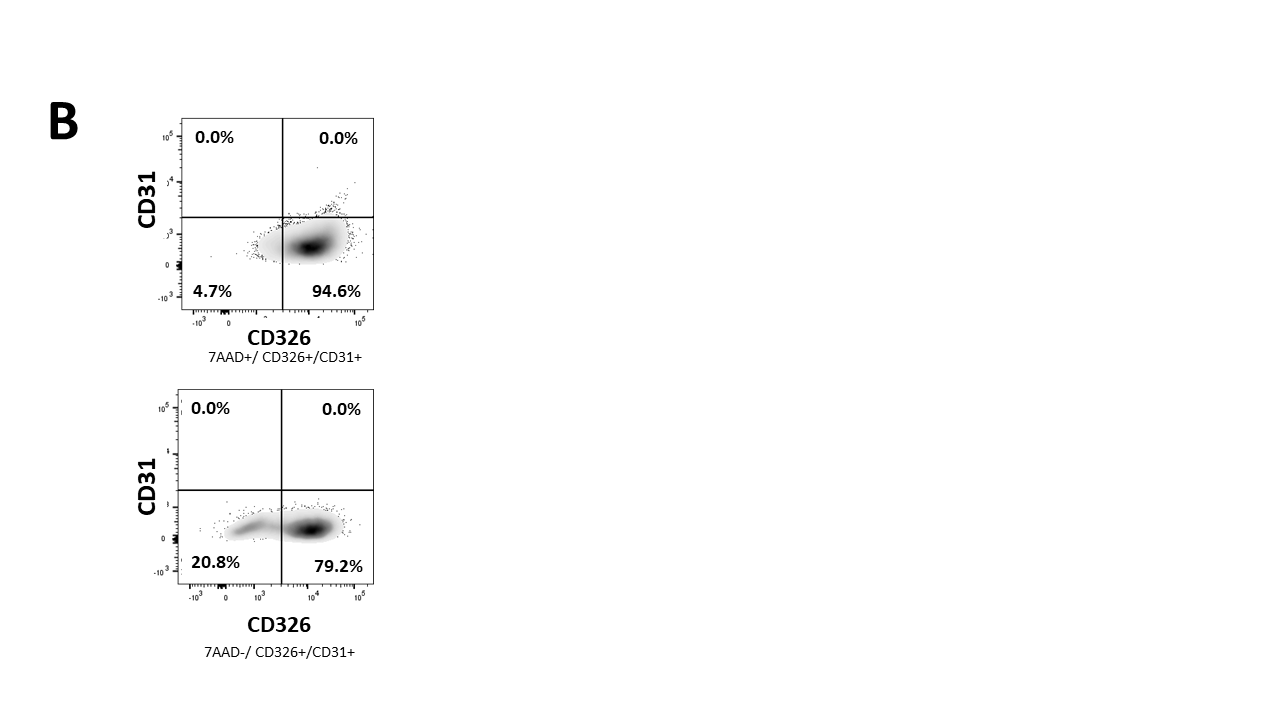

### Fig. S1C

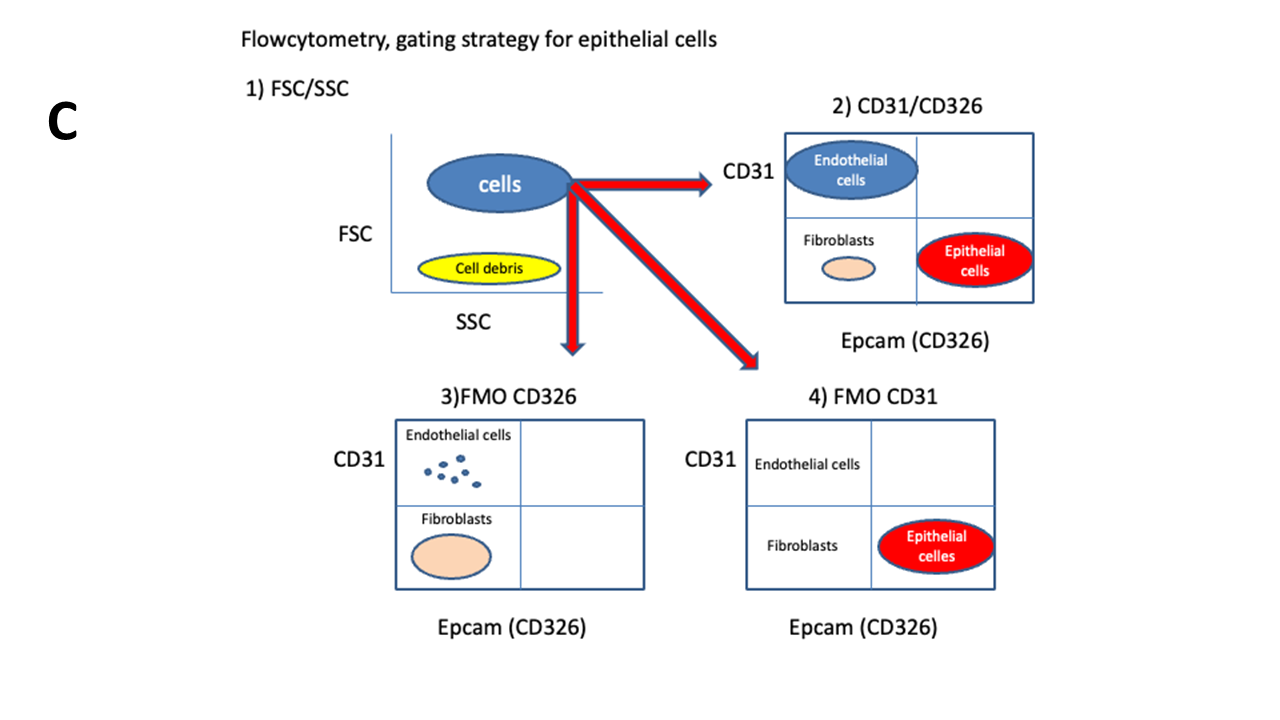

### Fig. S2A

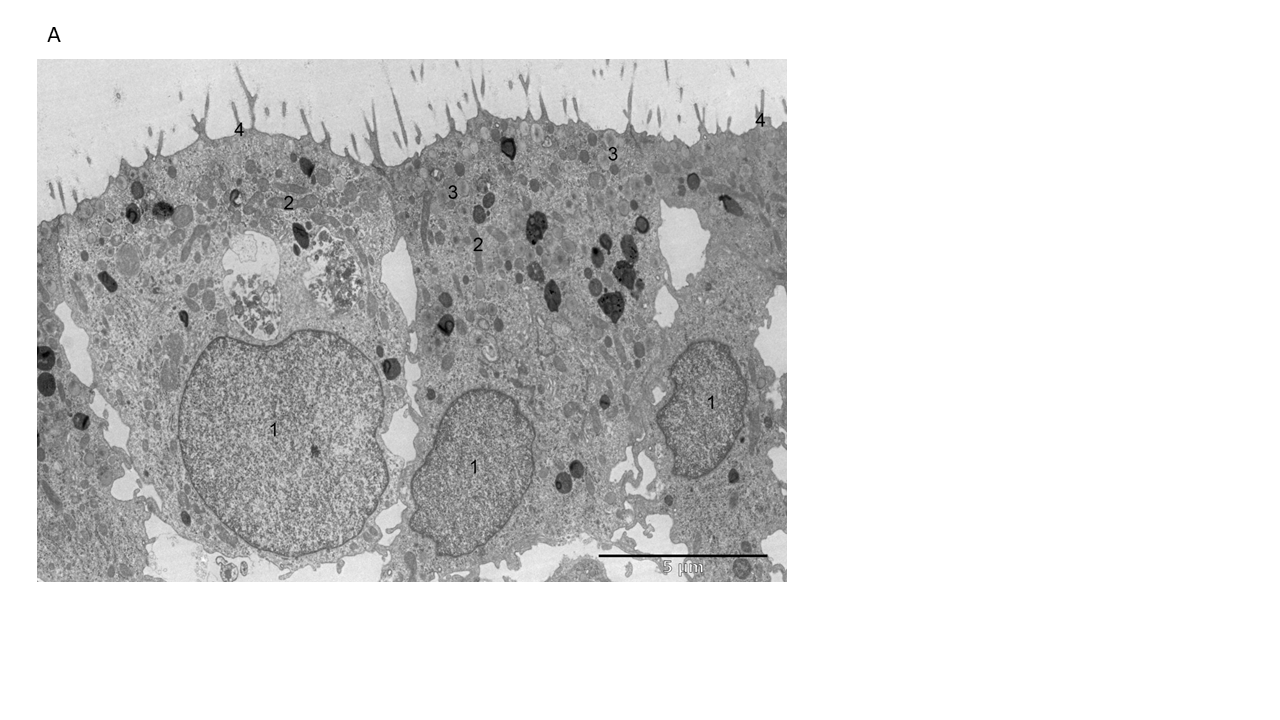

### Fig. S2B

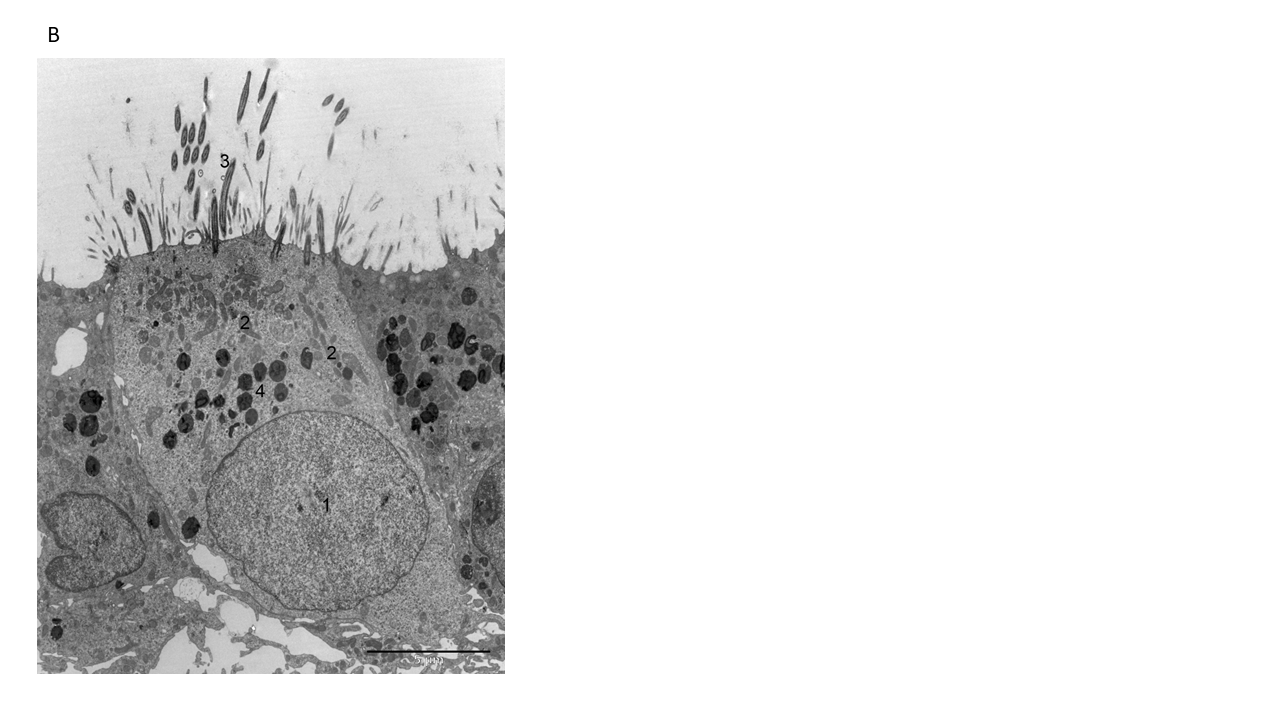

### Fig. S2C

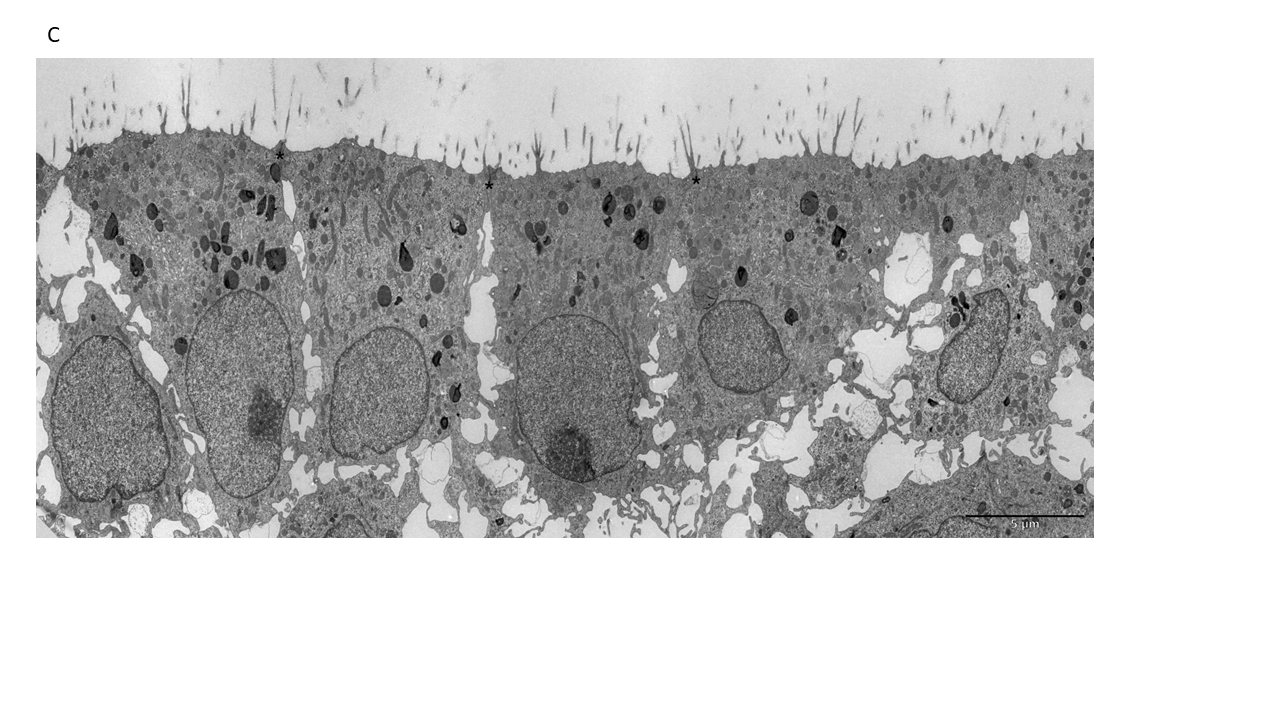

### Fig. S2D

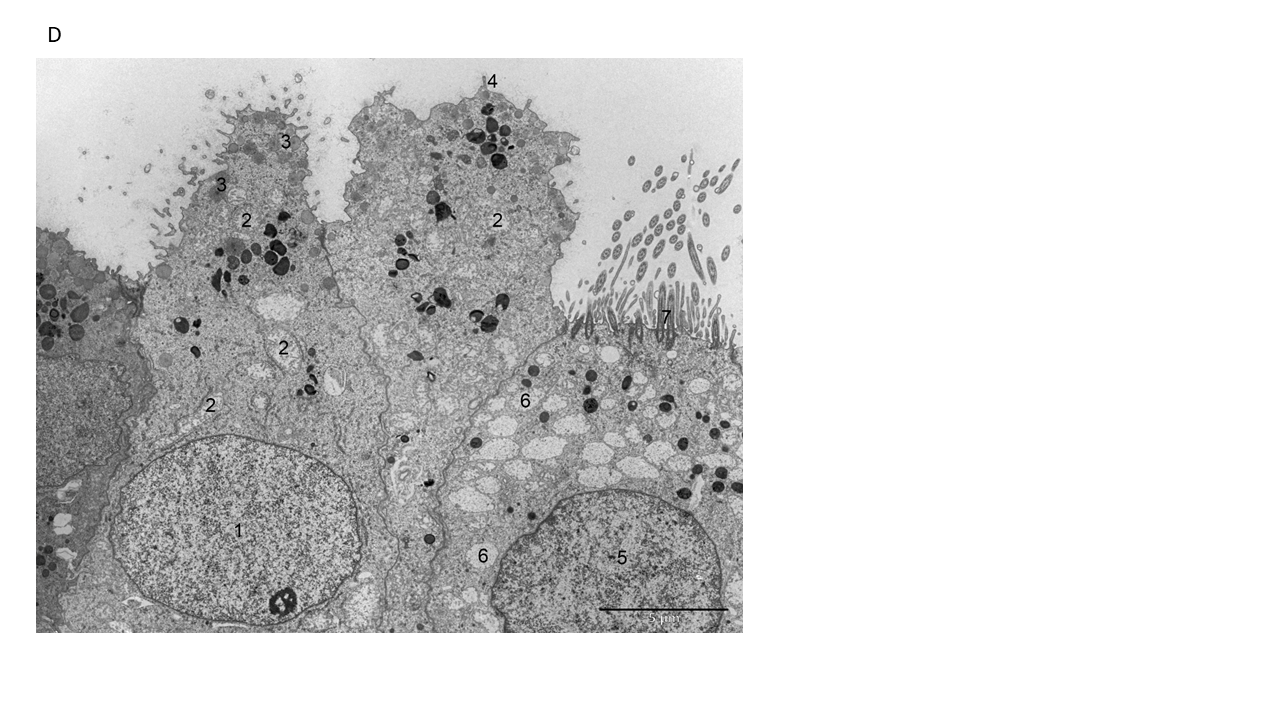

### Fig. S2E

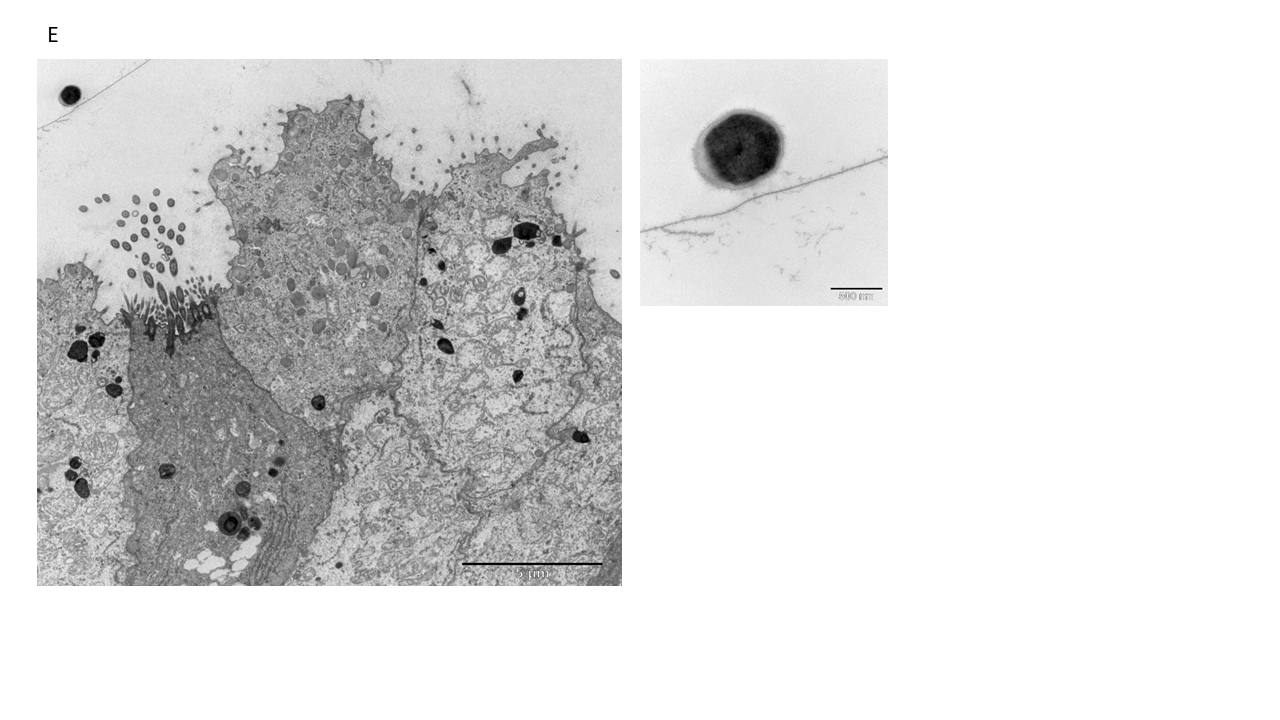

### Fig. S3

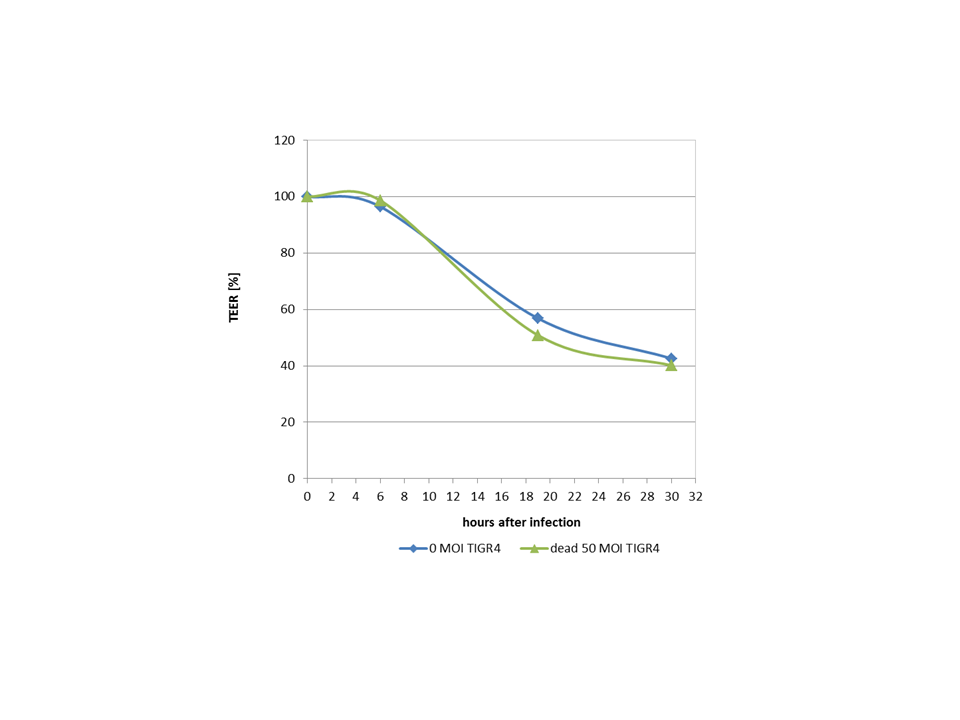
